# A new approach to recognize term and preterm infants with impaired kidney function (IKF) during the first week of life

**DOI:** 10.1101/2020.05.02.20089037

**Authors:** Sofia Perazzo, Mary Revenis, An Massaro, Billie L. Short, Patricio E. Ray

**Affiliations:** Division of Neonatology, The George Washington University, Washington DC 20010; Center for Genetic Medicine Research, Children’s National Hospital, The George Washington University, Washington DC 20010; Department of Pediatrics, The George Washington University, Washington DC 20010; Child Health Research Center, Department of Pediatrics University of Virginia School of Medicine, Charlottesville, VA 22908

**Keywords:** Neonatal AKI, renal function, first week of life, term and preterm newborns

## Abstract

**Background:** Current definitions of neonatal acute kidney injury (nAKI) are not sensitive enough to identify all newborns with impaired kidney function (IKF) during the first week of life. Previous studies in term newborns with hypoxic ischemic encephalopathy showed that the rate of serum creatinine (SCr) decline during the first week of life could be used to assess their renal status.

**Methods:** We reviewed the medical records of 329 critically ill newborns ≥ 27 weeks of gestational age (GA), to determine whether the rate of SCr decline combined with SCr thresholds provides a sensitive approach to identify newborns with IKF during the first week of life.

**Results:** Excluding neonates with nAKI, identified based on standard definitions, a SCr decline < 31 % by the 7^th^ day of life, combined with a SCr threshold ≥ 0.7 mg/dl, recognized newborns of 40-31 weeks of GA with IKF. A SCr decline < 21% combined with a SCr threshold ≥ 0.8 mg/dl identified newborns of 30-27 weeks of GA with IKF. These neonates (~ 17%) showed a more prolonged hospital stay and required more days of mechanical ventilation, vasoactive drugs, and diuretics, when compared to critically ill controls. Changes in urine output did not distinguish newborns with IKF.

**Conclusion:** The rate of SCr decline combined with SCr thresholds identifies newborns with IKF during the first week of life. This distinctive group of newborns that is missed by standard definitions of nAKI, warrants close monitoring in the NICU to prevent acute and chronic renal complications.

## Introduction

Critically ill newborns are at high risk of developing acute kidney injury (AKI) due to multiple factors, including intravascular volume depletion, capillary leak, ischemia, low cardiac output, nephrotoxic medications, and multiple organ dysfunction [1]. Under these circumstances, AKI is defined by a sudden decrease in kidney function, and is best diagnosed by measuring changes in the glomerular filtration rate (GFR) [2, 3]. However, due to the practical challenges involved in measuring the GFR in newborns, the diagnosis of AKI is made by assessing changes in serum creatinine (SCr), estimated creatinine clearance (CrCl), and urine output [4–20] Nonetheless, establishing a diagnosis of AKI on neonates can be problematic, in particular, during the first week of life, when there is no baseline steady-state SCr. In addition, the formulas used to estimate CrCl have not been validated in newborns [21–23], and changes in urine output are less reliable in neonates with low urinary concentration ability [12, 24]. All these limitations highlight the need to develop a more sensitive approach to assess the renal status of newborns during the first week of life.

Despite multiple studies, the development of a standardized definition of neonatal AKI remains a focus of active investigation and controversies [5, 25]. In particular, the first days of life create a unique dilemma. Due to the transfer of maternal SCr, waiting for a rise in SCr of the neonate may delay the recognition of the early stages of AKI. Additionally, the well-recognized limitations of SCr to assess the renal function in older children and adults, which are based on the observation that at higher GFR the SCr does not change rapidly [26], do not apply during the first week of life. Given the low GFR values of newborns (< 35 ml/min/1.73 m^2^) [2, 3], small changes in GFR could be associated with an exponential increase in SCr, as reported in adults with low GFR values [26] (Figure 1).

**Figure legend 1.**
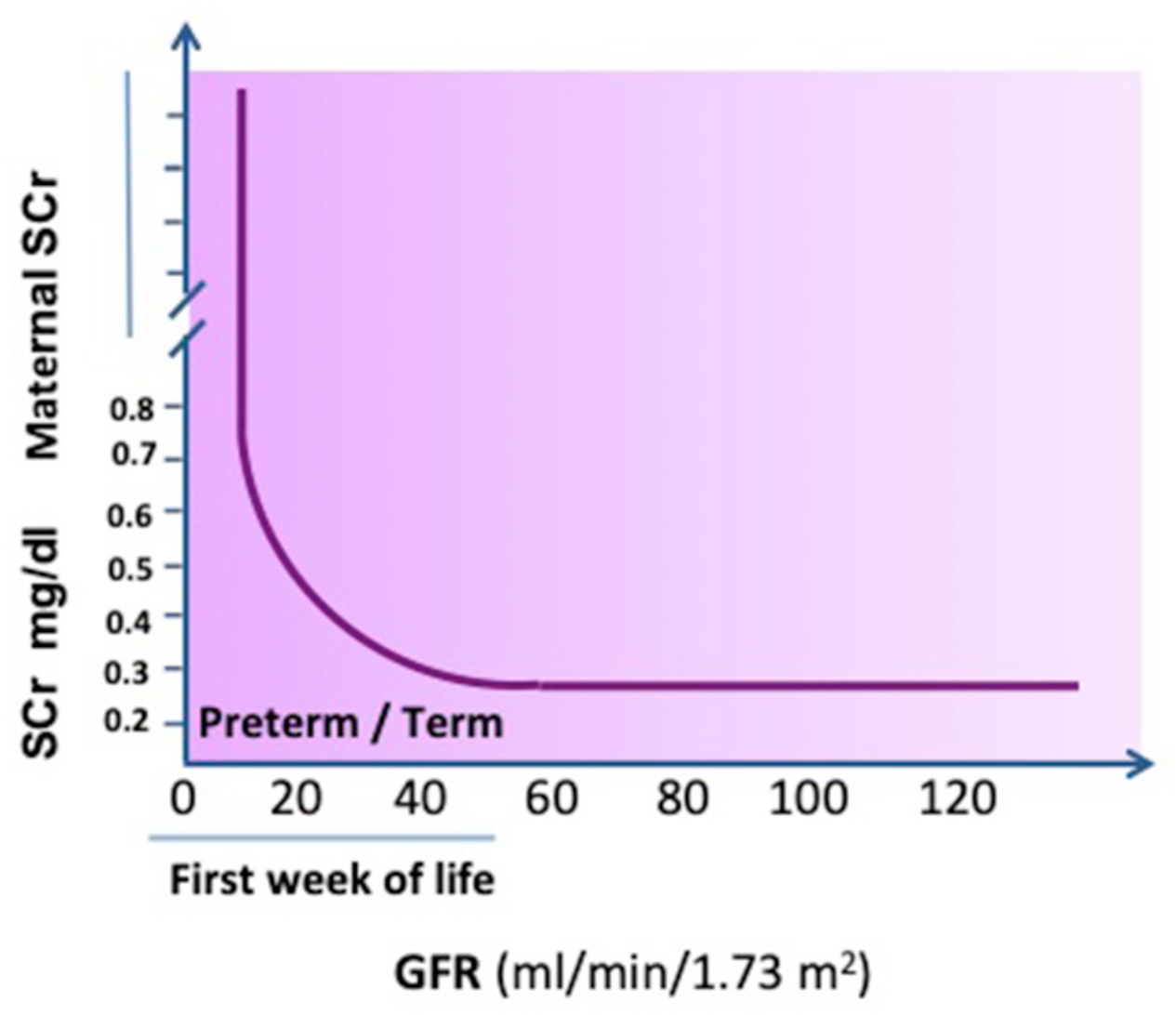
*Expected changes in SCr levels based on GFR* status (graph adapted from refs 6 and 26). The panel shows that at higher levels of kidney function (higher GFR values), large changes in GFR result in small or no changes in SCr. In contrast, at lower levels of kidney function (GFR < 40 ml/min/1.73 m^2^), small changes in GFR result in larger changes in SCr. Term and Preterm newborns have low GFR values during the 1^st^ week of life,

In a previous study we reviewed the outcome of 106 near-term newborns with hypoxic ischemic encephalopathy (HIE) treated with hypothermia during the first week of life [1]. We found that a standard AKI definition derived from the Kidney Diseases Improving Global Outcomes (KDIGO) guidelines, missed ~ 20 % of newborns who showed a significant delay in the rate of decline of SCr, and developed clinical complications associated with AKI. These patients required more days of mechanical ventilation and vasopressor drugs, had higher gentamicin levels, more fluid overload, lower urinary epidermal growth factor levels, and a more prolonged length of hospital stay, when compared to control neonates with HIE treated in a similar manner. Thus, it appears that the KDIGO-AKI definition is not sufficiently sensitive to detect all neonates who develop clinical complications associated with AKI events.

We carried out this study to validate and expand our previous findings, and determine whether the rate of SCr decline combined with thresholds for SCr, could be used to assess the renal status of term and preterm newborns with different critical illnesses during the first week of life,

## Material and Methods

### Study population

We conducted a retrospective review of 981 medical records of term and preterm neonates treated for a variety of diseases in a level 4 neonatal intensive care unit (NICU) at Children’s National Hospital, in Washington DC. Unidentified data were collected using the Research Electronic Data Capture (REDCap) database hosted in our institution between July 2011 and March 2015 [27]. Entry criteria included all term and preterm newborns (≥27 weeks of GA) who were admitted, survived the first week of life, and had at least one serum creatinine (SCr) measured during the first 48 hours of life and the 7^th^ day of life. Whenever the SCr levels were unavailable on day 7^th^, we included patients with SCr levels obtained on days 6 or 8 of life. Three hundred and twenty nine patients met these entry criteria and their medical records were reviewed. The study was approved by the IRB from the Children’s National Hospital and was conducted following the ethical guidelines of our institution with a waiver of consent.

### Definition of the study groups and selection of control patients

Newborns were divided in four groups according to their GA and mean SCr levels at the end of the first week of life. These groups included term infants of –37-40 weeks of GA, as well as pre-term infants of 34-36, 31-33, and 27-30 weeks of GA respectively. As described in previous studies for term infants, we defined the normal SCr values on day of life 7 as those within the 97.5th percentile (0.6 mg/dl) or lower, which is approximately two standard deviations (0.2 mg/dl) above the mean SCr levels of normal newborns [6, 28]. To define the cutoff SCr values for all other preterm groups, we calculated the mean SCr levels excluding all patients with AKI based on the KDIGO definition, and defined the abnormal threshold at 0.2 mg/dl above the mean. By using this approach, we took into consideration the standard error of the SCr measurements, as well as an adjustment for the normal fluid losses expected during the first week of life [29]. The control neonates in each group included all newborns with normal SCr values at the end of the first week of life. The impaired kidney function (IKF) group included all newborns with abnormal SCr levels at the end of the first week of life, excluding those in the AKI-KDIGO group. Neonates who showed abnormal SCr levels during the 3^rd^ and 6^th^ days of life, whenever available, were assigned to the IKF group, unless they met the criteria to enter the AKI-KDIGO group. The AKI-KDIGO group included newborns who developed AKI according to the standard neonatal KDIGO definition, based on the rise of SCr ≥ 0.3 mg/dl within any 48 hr. period [30].

### Serum Creatinine decline

The rate of SCr decline was calculated by estimating the % SCr decline from the first 48 hours of life, which represents the transfer of maternal SCr, until day of life 7. We then performed ROC analysis to establish the cutoff points for the SCr decline in the control and IKF groups. Newborns showing SCr values during the first 48 hours of life within the normal range for the 7^th^ day of life, which suggest a limited transfer of maternal SCr, were excluded from this analysis. In these cases, further changes in SCr below the normal levels for the first week of life have limited clinical value.

### Data and sample collection

All relevant demographic and clinical data were recorded including mortality, days of hospital stay until disposition, use and duration of mechanical ventilation, vasoactive drugs, and diuretics. Urine output was measured according to the NICU standard protocol of care [6]. SCr was measured at Children’s National Central laboratory using either the Jaffe or enzymatic methods.

### Statistical analysis

Demographic and clinical data are reported as the mean ± SD for continuous variables and as proportions. All data sets were tested for normality with the D’Agostino and Pearson omnibus normality test. The median with interquartile range (IQR) was used for non-normally distributed variables. Differences between two groups were analyzed by the Student t test or Mann–Whitney U tests where appropriate. Proportional differences were tested with the Fisher exact test or the Chi-square test with Bonferroni’s correction, whenever indicated. Intergroup comparisons were tested with analyses of variance (ANOVA). Receiver-operating characteristic (ROC) curves were used to define the SCr cutoff points for optimal sensitivity and specificity to identify newborns with IKF. The statistical analysis was done using the Prism 5 and MedCal software programs from GraphPad Software, Inc., La Jolla, CA, and Oostend, Belgium respectively.

## Results

### Demographics and clinical characteristics

We examined 981 medical records of newborns who were admitted with a variety of diagnoses, including sepsis, respiratory distress syndrome, hypoxic ischemic encephalopathy, cardiac malformations, intraventricular hemorrhage, prematurity, surgical conditions, and other illnesses. From this group, 329 newborns met the study entry criteria, and their medical records were reviewed in depth. The baseline birth weight, gender, and race, are shown in Table 1. The birth weight in grams (mean ± SD) of the newborns in each of the corresponding GA group were: 3089 ± 666 (37-40 weeks of GA); 2514 ±547 (34-36 weeks of GA); 1714 ± 424 (31-33 weeks of GA); and 1239 ± 348 (27-30 weeks of GA).

**Table 1.**
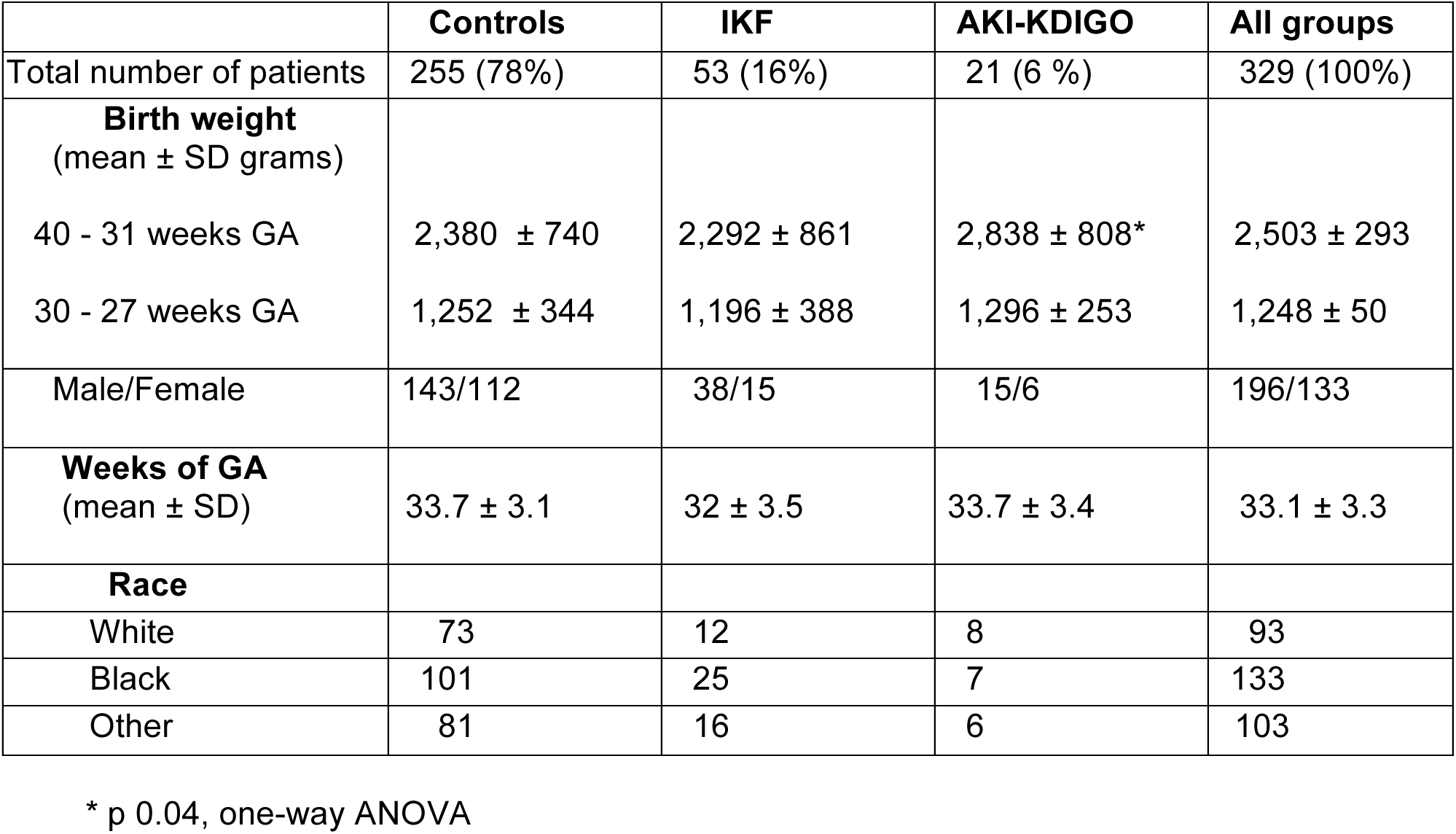
Demographic characteristic of the newborn study groups

|  | **Controls** | **IKF** | **AKI-KDIGO** | **All groups** |
| --- | --- | --- | --- | --- |
| Total number of patients | 255 (78%) | 53 (16%) | 21 (6 %) | 329 (100%) |
| **Birth weight**<br>(mean ± SD grams) | | | | |
| 40 - 31 weeks GA | 2,380 ± 740 | 2,292 ± 861 | 2,838 ± 808* | 2,503 ± 293 |
| 30 - 27 weeks GA | 1,252 ± 344 | 1,196 ± 388 | 1,296 ± 253 | 1,248 ± 50 |
| Male/Female | 143/112 | 38/15 | 15/6 | 196/133 |
| **Weeks of GA**<br>(mean ± SD) | 33.7 ± 3.1 | 32 ± 3.5 | 33.7 ± 3.4 | 33.1 ± 3.3 |
| **Race** |  |  |  |  |
| White | 73 | 12 | 8 | 93 |
| Black | 101 | 25 | 7 | 133 |
| Other | 81 | 16 | 6 | 103 |

\* p 0.04, one-way ANOVA

### Renal status, creatinine levels, and rate of serum creatinine decline

Changes in SCr decline and absolute values are shown in Figures 2–3, and table 2. The SCr levels were elevated on the first 48 hours of life, reflecting the transfer of maternal SCr. No significant differences in SCr levels were noted between the different GA groups at this time point. Approximately 7% of the neonates developed AKI based on the KDIGO definition, and these patients showed the highest SCr levels by the end of the first week of life (2.6 ± 1.9 mg/dl; mean ± SD). No significant differences in SCr decline or absolute SCr values were found between term infants (37-40 weeks GA) and late preterm infants (34-36 GA) (p= 0.05) (Figure 3). Moderate preterm infants (31-33 weeks of GA) showed a modest but significant increase in SCr levels (p = 0.02), but no significant differences in the SCr decline values (> 0.05) when compared to the 34-36 GA group (Figure 3). As expected, very preterm infants (30-27 weeks of GA) had the lowest SCr decline (p < 0.01) and the highest absolute SCr levels (p < 0.001) when compared to all other groups (Figure 3). Based on these data, we established the absolute SCr abnormal cutoff values for the end of the first week of life, and used these values to divide the patients in two groups for further analysis. These groups were newborns of 40 - 31 weeks of GA (SCr cut-off ≥ 0.7 mg/dl), and newborns of 30-27 weeks of GA (SCr cut-off ≥ 0.8 mg/dl).

**Figure legend 2.**
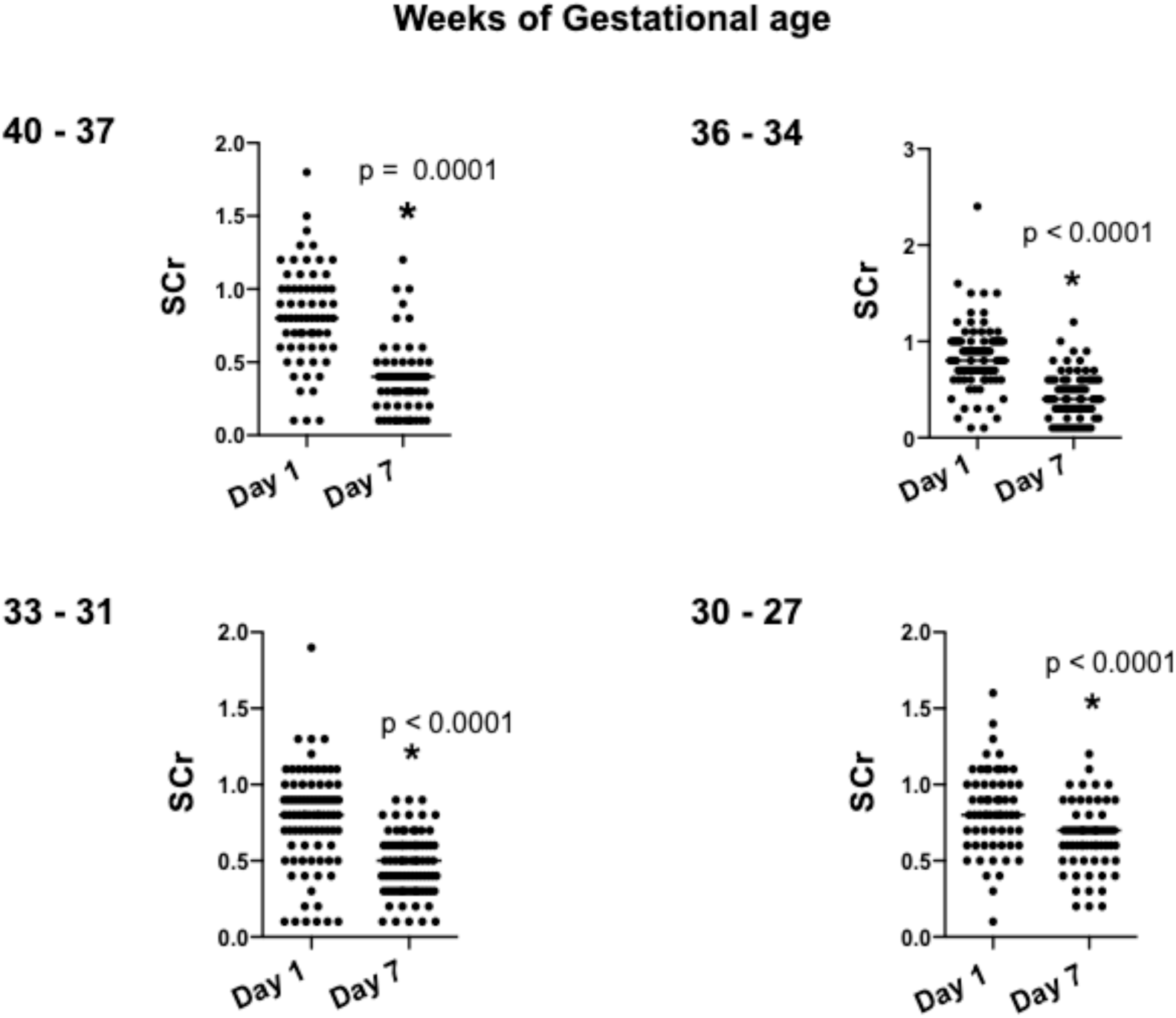
SCr levels during the first week of life in neonates of different gestational ages. The panels show representative SCr values for the first and seven days of life in neonates corresponding to each of the four gestational age groups, excluding patients who developed AKI according to the KDIGO definition. ^*^ P values < 0.05 were considered statistically significantly by unpaired t test.

**Figure Legend 3.**
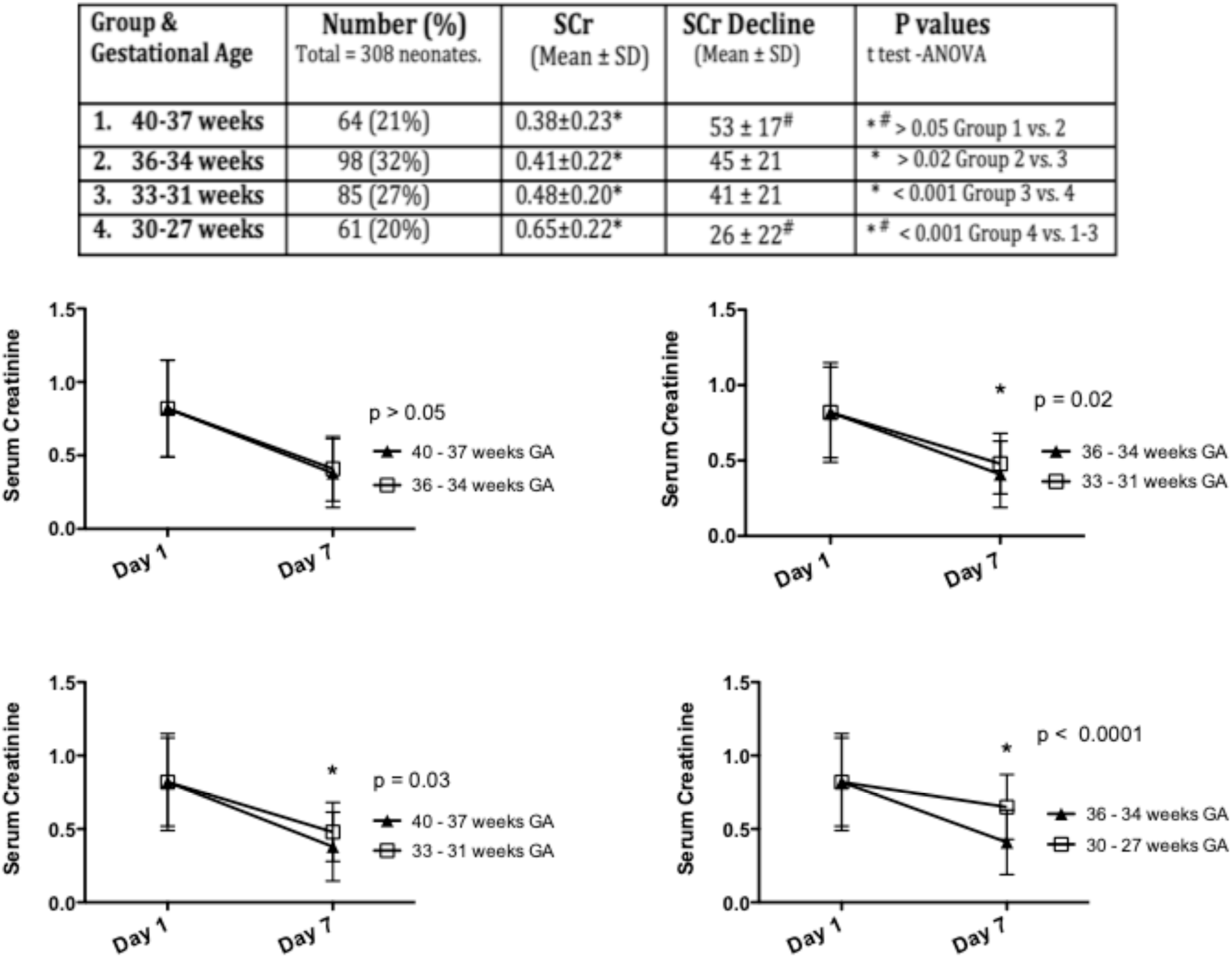
Rate of decline of SCr in newborns during the first week of life. The upper panel shows the total number of newborns in each group, as well as the SCr, and SCr decline values in each group. The graphs below compare the rate of decline of SCr between groups of different gestational ages. P values < 0.05 were considered statistically significant by unpaired t test.

**Table 2.**
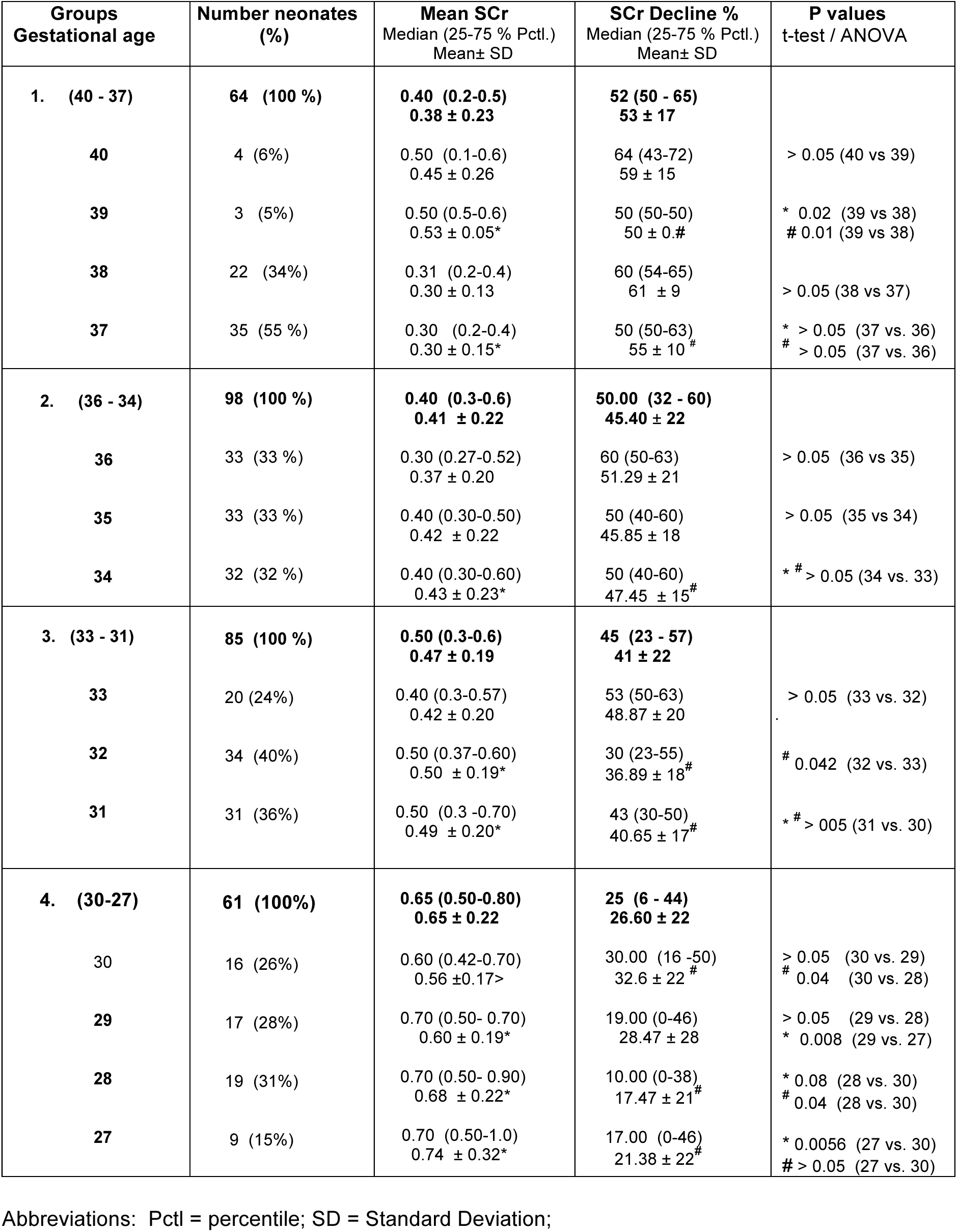
Mean SCr and SCr decline values in critically ill newborns during the 1^st^ week of life

ROC curve analyses of the SCr decline including eligible controls and IKF neonates are shown in Figure 4. When all newborns of 40 to 27 weeks of GA were analyzed together, using the corresponding SCr cut-off points for each group, a SCr decline criterion of <31%, yielded ~ 81% sensitivity, and 86% specificity to discriminate all newborns of –27-40 weeks of GA with IKF (AUC 0.92; p < 0.001) (Fig. 4A). ROC curve analyses of newborns –31-40 weeks of GA yielded a sensitivity 84% and specificity 96%, to discriminate IKF (AUC 0.925; p < 0.001). Preterm infants 27-30 weeks of GA yielded significant lower SCr decline values (Fig. 4B), and a higher SCr cutoff point by the end of the first week of life (≥ 0.8 mg/dl). Overall, a criterion < 21% showed ~ 93 % sensitivity and 93% specificity to discriminate preterm infants of 30-27 weeks of GA with IKF (AUC 0.941; p < 0.001).

**Figure Legend 4.**
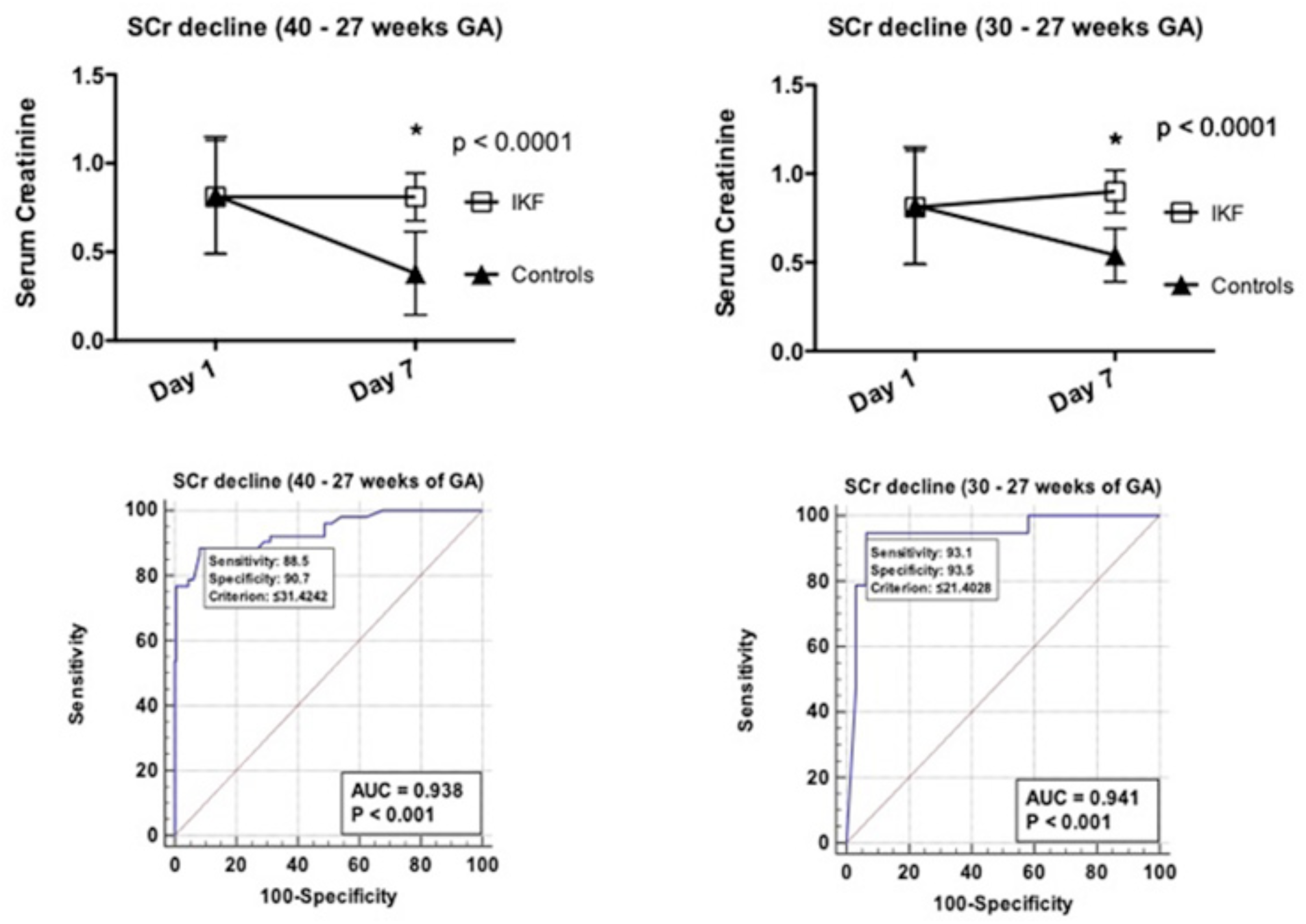
Rate of SCr decline in newborns with IKF. The upper panels show the rate of SCr decline values for newborns with IKF compared to the controls of 40 −27 weeks of GA and 30 to 27 week of GA. All newborns with AKI were excluded from this analysis, ^*^ P < 0.05 were considered statistically significant by an unpaired t test. The lower panels show receiving operational characteristic (ROC) curves generated from data derived from both groups, to define the SCr decline cut-off values at day 7^th^ of life. The cutoff values and predicted ability of SCr to detect newborns with IKF was determined excluding all patients who developed AKI based on the KDIGO definition. The area under the curve (AUC) for each group is displayed on the lower panels. The dashed reference line represents a ROC curve for a test with not discriminatory ability. The sensitivity, specificity and criterion values are shown on the lower panels. P < 0.05 were considered statistically significant

### Clinical outcome

When compared to the control group, neonates with IKF had higher mortality rates, a more prolonged hospital stay, and a greater requirement for mechanical ventilation, vasoactive drugs, and diuretics during the first weeks of life (Figure 4). The BUN levels were significantly higher in patients with IKF when compared to controls, although neonates with AKI showed the highest BUN levels (Figs 5–6). No significant differences in urine output were noted between control and IKF neonates (Fig. 6). In contrast, newborns in the AKI-KDIGO group showed more significant oliguria, need of diuretics, and higher mortality rates, when compared to all other groups (Fig 6). No statistical significant differences in the total bilirubin levels were noted between control and IKF newborns by the end of the 1^st^ week of life. Briefly, the bilirubin plasma levels were 7.9 ± 4.2 and 7.6 ± 4.8 mg/d in the controls and IKF newborn groups of 31-40 weeks of GA respectively, and 4.8 ± 1.8 and. 5.2 ± 2.8 mg/dl for newborns of 27-30 weeks of GA in the control and IKF groups respectively.

**Figure Legend 5.**
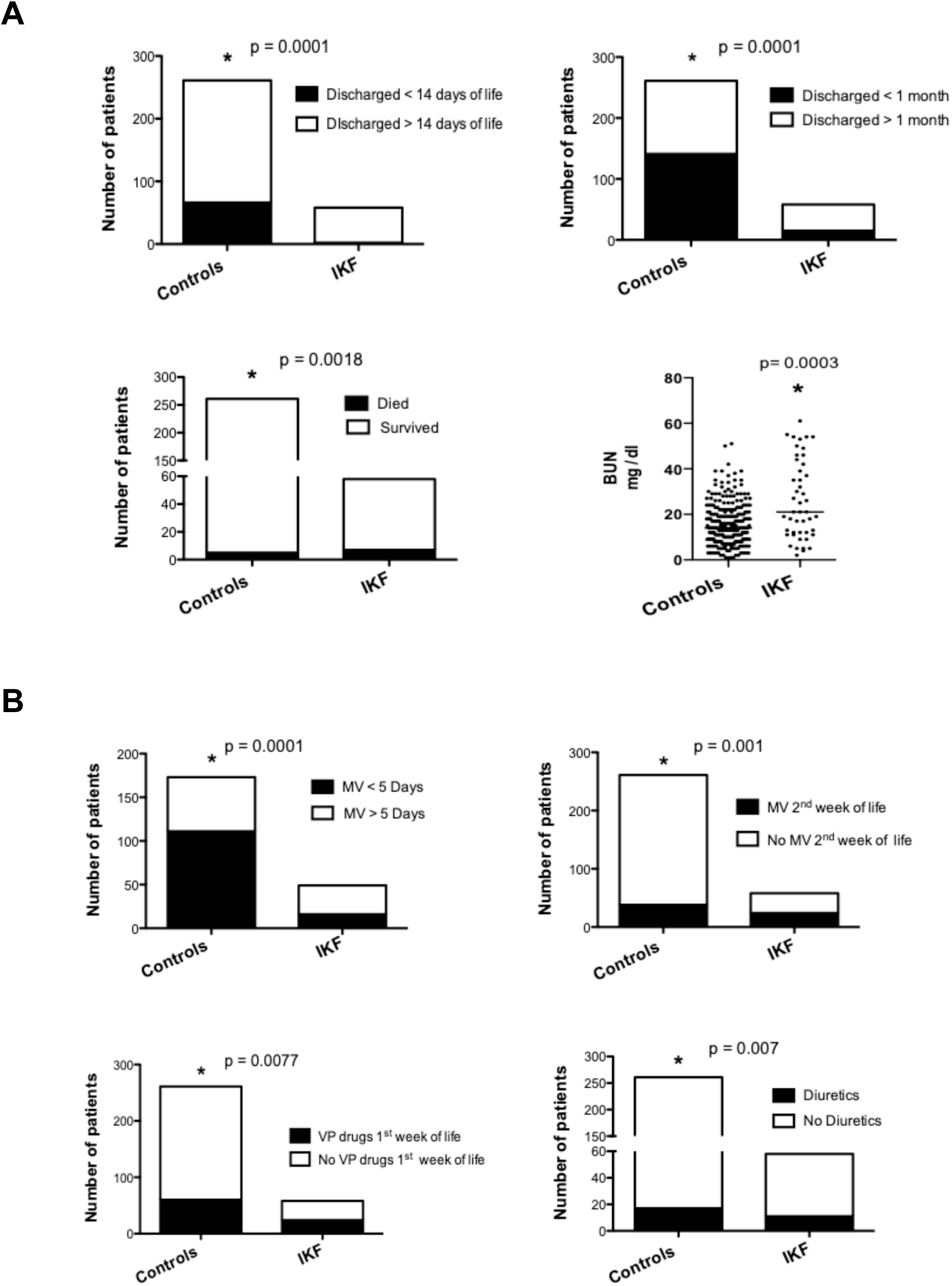
Clinical outcome of newborns with IKF. (A) Patients in the IKF group were discharged after a more prolonged hospital stay, and had higher mortality rates, and Blood Urea Nitrogen levels, when compared to the control group. (B) Patients in the IKF group required more days of mechanical ventilation (MV), more vasopressor (VP) drugs, and more diuretics during the first two weeks of life, when compared to controls. All patients with AKI according to the KDIGO definition were excluded from this analysis.

**Figure legend 6.**
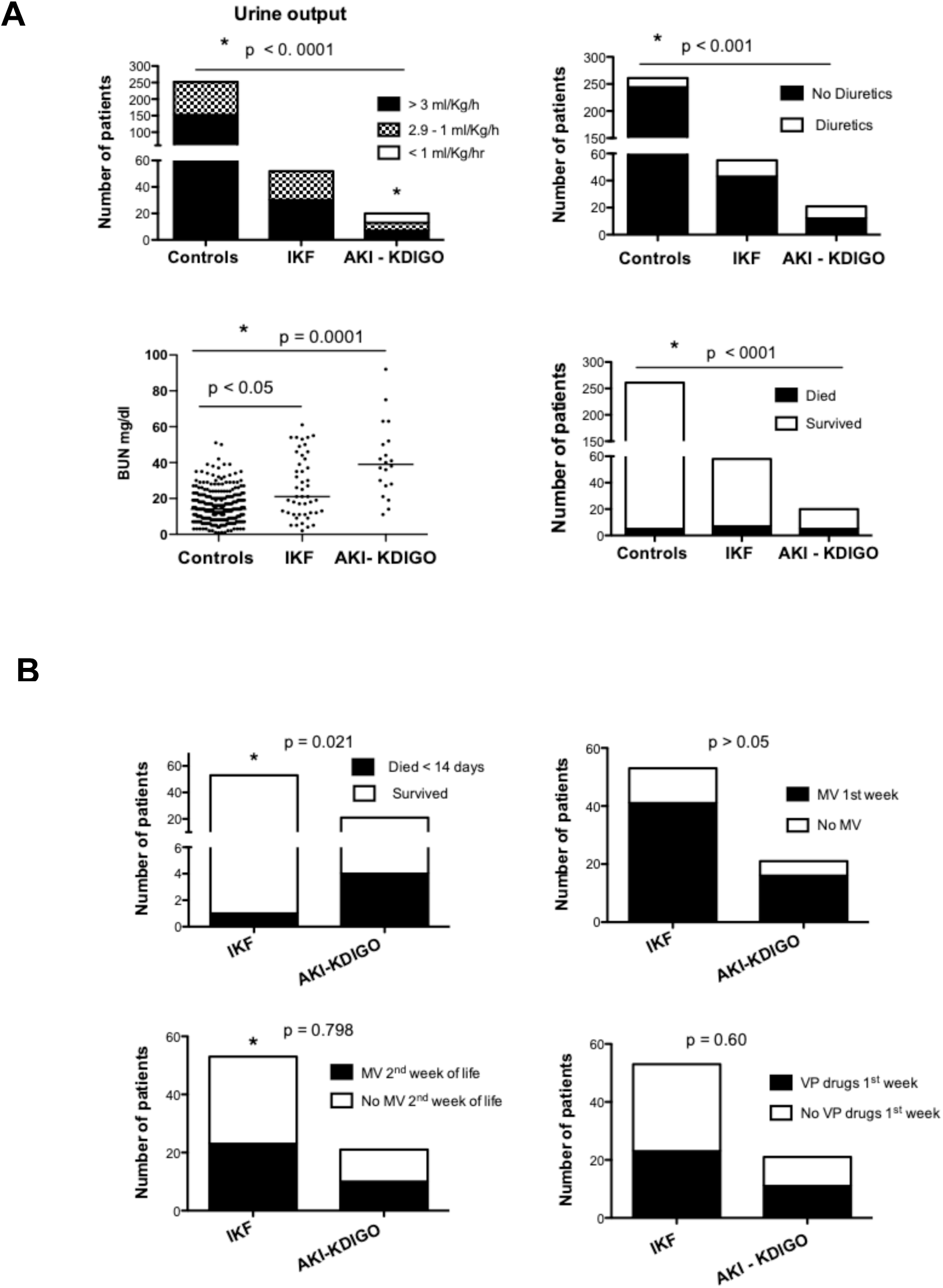
Clinical outcome of newborns with impaired kidney function (IKF) and AKI. (A) Newborns with AKI according the neonatal KDIGO definition (AKI-KDIGO) showed more oliguria, higher BUN levels, required more diuretics, and showed higher mortality, when compared to the control and IKF groups. ^*^ P < 0.05 was considered significant by ANOVA. (B) Patients in the AKI-KDIGO group showed higher mortality during the first two weeks of life. No significant differences in days needed of mechanical ventilation (MV) or vasopressor (VP) drugs were noted between patients with IKF and AKI-KDIGO. ^*^ P < 0.05 was considered statistically significant by unpaired t test.

## Discussion

The major finding of this study is that a rate of SCr decline < 31% combined with a threshold for SCr ≥ 0.7 mg/dl by the end of the first week of life, provide a new and sensitive approach to identify term and preterm infants >30 weeks of GA with impaired kidney function (IKF). We used the new term IKF because changes in SCr decline and absolute SCr levels during the first week of life reflect changes in kidney function that may not be necessarily associated with an acute kidney injury event. Moreover, our data suggest that the standard neonatal AKI-KDIGO definition is not sufficiently sensitive to identify all newborns with IKF during the first week of life. We propose that neonates showing abnormal SCr threshold and decline values by the end of first week of life constitute a distinctive clinical group of newborns with IKF that warrants close monitoring in the NICU to prevent acute and chronic renal complications.

To the best of our knowledge, this is the first study to use SCr decline values combined with absolute SCr cutoff points to assess the renal status of preterm infants during the first week of life. In this manner, we were able to identify a group of newborns with IKF, who were missed by the AKI-KDIGO definition, and required more days of mechanical ventilation, more continuous use of vasopressor drugs, diuretics, and had an increased length of hospital stay when compared to the control group of critically ill newborns. Newborns with AKI based on the KDIGO definition, showed similar clinical complications, although more severe oliguria, increased need of diuretics, and higher mortality rates. Although diuretics can mask the detection of oliguria as an early sign of AKI, our findings are in agreement with previous studies showing that oliguria is not a sensitive clinical sign of neonatal AKI [11, 12, 24]. Taken together, our findings suggest that the rate of SCr decline combined with SCr thresholds provide a sensitive approach to assess the renal function of newborns during the first week of life. More studies are warranted to assess the short and long-term clinical and renal complications of neonates who develop IKF during the first week of life.

As expected, pre-term infants of 27-30 weeks of GA showed the lowest rate of SCr decline and highest SCr levels during the first week of life [31]. In humans, the process of nephrogenesis is completed between 32-35 weeks of GA [5, 32]. Thus, the rate of SCr decline appears to correlate with the renal physiological and maturational changes characteristic of preterm infants during this time. In agreement with this notion, the SCr levels at the end of the first week of life were higher in control newborns of ~ 32 week of GA, when compared to term and near-term neonates. Subsequently, the baseline normal SCr levels, further increased in control neonates of 30 and 27 weeks of GA respectively, probably reflecting different stages of renal development and maturation at these time points. Interestingly, we did not see a rise in SCr during the first days of life in control newborns of 27-30 weeks of GA, as reported in previous studies. In contrast, we were able to generate SCr decline and absolute SCr cutoff points for this group as well. However, given the limited number of newborns in this group, our results need to be validated in a larger cohort. Nonetheless, our data suggest that the SCr decline could be used in a reliable manner to assess the renal status of preterm neonates > 30 weeks of GA.

In addition to the KDIGO definition, neonatal AKI has been defined by other methods [33], including a SCr greater than 1.5 mg/dl or an increase of at least 0.2 to 0.3 mg/dl per day from a previous lower value. Several SCr AKI cutoff points have been recommended for preterm infants between 24 to 32 weeks of GA (> 1.6, > 1.1 and > 1.0 SCr mg/dl, for infants of 24 to 27, 28 to 29, and 30 to 32 weeks of GA respectively) [1]. These cutoff points, however, have not been validated during the first week of life, and if they are applied to our cohort, they will miss a significant number of newborns with IKF. The rationale to focus our approach on the SCr decline is based on the transfer of maternal SCr that occur after birth, and the changes associated with the maturation of renal function. During the first few hours of life, the GFR reflects the functional renal status in utero, when the placenta plays a key role in this process. Subsequently, during the first 3 days of life, there is a rapid increase in renal function [11, 28, 32, 34, 35], and given the low GFR values of newborns [20], the SCr can change in an exponential fashion in response to a small change in GFR, as reported in adults with GFR levels < 40 ml/min/1.73m^2^ [26]. Nonetheless, due to the transfer of maternal SCr, a significant renal injury event may not necessarily increase the SCr levels by 0.2 or 0.3 mg/dl. Our previous study in term newborns with hypoxic ischemic encephalopathy (HIE) supports this notion, and shows that the neonatal AKI-KDIGO definition misses a significant number of newborns undergoing the early stages of AKI. The current study validated and expanded our previous results in term and preterm infants affected with different illnesses.

Previous studies have used creatinine (Cr) kinetic models to review the definitions of AKI in adults [36]. These models are based on the mass balance principles, and predict that to induce a percent rise of SCr by 50%, more Cr is accumulated in people with higher baseline SCr levels compared to those with normal baseline levels. Thus, assuming a constant Cr generation rate and volume of distribution, the changes in GFR levels in people with high baseline SCr levels, can be estimated in a more precise manner by considering absolute changes in SCr rather that a % rise [36]. Furthermore, the time to reach a 50% increase in SCr is shorter in patients with lower SCr baseline values [36]. During the first week of life, the renal status of newborns could be compared to people with chronic kidney disease. Both show low GFR values and higher SCr levels relative to the body mass and Cr generation rates. It could be argued that theoretical calculations based on the mass balance principles cannot be applied to the first week of life, since the SCr is not in a steady-state balance and its volume of distribution it’s changing. Nonetheless, assuming that these variables follow the same pattern in newborns of similar GA, the kinetic models underscore the limitations of using a % rise in SCr to estimate changes in GFR during the first week of life. For this reason, we established absolute SCr cutoff point to interpret the changes in SCr decline. In addition, to establish the SCr threshold, we took into consideration the standard error of the SCr measurements, as well as an adjustment for the normal fluid looses that occur during the first week of life. Finally, we also noted that the rate of SCr decline has additional value to assess the renal status of neonates born with abnormal SCr levels (≥ 1.2 mg/dl). In fact, ~ 60% of the neonates who showed normal SCr decline values associated with abnormal SCr levels by the end of the first week of life, had favorable clinical outcomes. Therefore, both the SCr decline and thresholds values should be taken into consideration to assess the renal status of newborns during this time.

We acknowledge that the ideal definition of AKI should rely on the discovery of specific biomarkers of kidney injury. Unfortunately, currently we lack specific biomarkers of renal injury for the first week of life [5, 37], and all neonatal AKI definitions are based on assessing % or absolute changes in SCr [5, 6, 19, 30, 37], which reflect changes in kidney function rather than injury. For this reason, we propose to use the term impaired kidney function (IKF) to group neonates with a slow SCr decline during the first week of life. We recognize that the term IKF will include neonates undergoing hemodynamic changes leading to poor renal perfusion, extracellular volume depletion, acute or chronic kidney injury, drug-induced nephrotoxicity, and / or renal immaturity. Nonetheless, all these factors contribute to precipitate or exacerbate AKI events. In addition, newborns undergoing the early stages of AKI usually meet the IKF criteria. Thus, the early recognition of newborns with IKF provides a great opportunity to discover and validate new biomarkers of AKI during the first week of life. We should mention also that definitions of AKI that place too much emphasis on predicting the worst outcomes (e.g. mortality), could miss the early stages of AK and delay its recognition. In summary, we need a standardize definition of neonatal AKI that predict clinical outcomes, but that also recognizes the early stages of AKI, in order to initiate early treatments and discover new biomarkers of renal injury.

Our study has several limitations, including its retrospective nature, single center, lack of GFR measurements and more accurate calculations of urine output and fluid overload. In addition, the clinical outcomes associated with the diagnosis of IKF could be considered non-specific changes associated with systemic illnesses. However, these complications are similar to those seen in neonates with AKI. In addition, because Children’s National Hospital laboratory used two different methods to measure the SCr during the time frame of the study, we took into consideration the standard error of both methods to define the normal cut-off points, as well as the total bilirubin plasma levels, which can affect the SCr measurements using the Jaffe method, but were not significantly different between the control and IKF groups of newborns respectively. Furthermore, our SCr values are consistent with the values reported by large multicenter studies that have used both methods [30]. Finally, the relative small number of newborns with IKF and AKI in each GA group prevented us from doing a powerful statistical analysis in each individual group. Nevertheless, when data derived from more than 300 newborns are taken together, they provide strong statistical evidence to support our conclusions.

In summary, we propose that a SCr decline < 31 % in combination with a threshold for SCr ≥ 0.7 mg/dl by the end of the first week of life, provides a sensitive method to assess the renal function of term and preterm newborns > 30 weeks of GA. Hopefully, this approach will facilitate the early detection and treatment of neonates with IKF during the first week of life, provide and opportunity to discovery new and more specific biomarkers of AKI, and identify neonates at higher risk of developing acute or chronic kidney complications in the NICU.

## Data Availability

All data available is presented in the manuscript submitted.

## Notes

### Competing Interest Statement

The authors have declared no competing interest.

### Funding Statement

This study was supported by the National Health Institutes R01 grants # HL102497 and # DK049419

